# ARE PEOPLE WITH FAMILY HISTORY OF PANCREATIC CANCER ENRICHED WITH GENETIC PROPENSITY FOR DIABETES MELLITUS?

**DOI:** 10.1101/2025.08.15.25333774

**Authors:** Simon Egyin, David W. Sosnowski, Albert E. Orhin, Dominique S. Michaud, Elizabeth A. Platz

**Author notes:** Funding: This research was conducted without any external funding support.

## Abstract

**Background:** Observational studies suggest that long-standing diabetes may be a cause of pancreatic cancer. To further support this observation, we aimed to determine whether the genetic risk of diabetes is associated with a family history of pancreatic cancer in persons without a personal history of pancreatic cancer. We hypothesized that if diabetes is a cause of pancreatic cancer, then persons with a family history of pancreatic cancer, reflecting both inheritance and lifestyle risk factors, will be enriched with a genetic propensity for diabetes. Our hypothesis assumes that the causal genes for diabetes and pancreatic cancer are not identical.

**Methods:** We conducted a cross-sectional analysis of 3,911 participants without a personal history of pancreatic cancer using data from the *All of Us* Research Program. Family history of pancreatic cancer was assessed through structured electronic health record reviews and self-reported questionnaires capturing first- and second-degree relatives’ cancer histories. A polygenic score (PGS) for type II diabetes was created using short-read whole genome sequencing (srWGS) data in *All of Us*. Logistic regression was used to test the association between family history (first degree relative) of pancreatic cancer and polygenic risk for diabetes, adjusting for relevant demographic and lifestyle variables.

**Results:** Mean age was 65.7 years, with 64.8% women and 83.4% White participants. Among the 3,911 participants,173 individuals (4.4%) had a family history of pancreatic cancer. Diabetes PGS was not associated with a higher OR of a family history of pancreatic cancer; OR for higher PGS was 0.67 (95% CI 0.45-0.98) before adjustment and 0.67 (95% CI 0.44-0.97) after adjustment. Restricting to participants ≤50 years among whom we expected a lower likelihood of survival bias, the association was attenuated (OR= 0.75, 95% CI 0.47- 1.18). In contrast, the OR was even more inverse in participants >50 years (OR= 0.60, 95% CI 0.38-0.94). Simulations further support the potential for survival bias.

**Conclusions:** Contrary to the hypothesis, persons with a family history of pancreatic cancer were not enriched with genetic risk of diabetes. The observed inverse association may be partially due to survival bias. This work points to complexities in the conduct of etiologic research using a cross-sectional design.

## Introduction

Pancreatic cancer ranks among the deadliest cancers, with a five-year survival rate of just 13%. ^1^ The vast majority (about 90%) of cases are pancreatic ductal adenocarcinoma, an especially aggressive and difficult-to-treat form. ^2^ Globally, pancreatic cancer cases and deaths are expected to rise sharply by 2045, according to the Global Cancer Observatory. ^3^ With low survival rates and no screening available, prevention through risk factor management is essential.

A small number of factors are associated with pancreatic cancer risk, including type II diabetes, which is especially important due to its growing global impact. ^4,5^ Diabetes mellitus affects a significant portion of the U.S. population, with approximately 10% of Americans, or over 38 million individuals, living with the condition. ^6^ Most of these cases, ranging from 90% to 95%, are classified as type II diabetes mellitus. While this form of diabetes mellitus has traditionally been associated with middle-aged and older adults, there is a concerning trend of increasing diagnoses among children, adolescents, and young adults.^6^

This increase in diabetes prevalence in the U.S. and globally is likely due to an increase in the prevalence of modifiable risk factors.^7^ Yet, there remains strong evidence supporting an underlying genetic predisposition. The genetic basis for developing type II diabetes mellitus is characterized as polygenic, with over 50 genes detected ^8^, and there is no evidence that a single gene fully explains the risk of type II diabetes mellitus, except in limited populations. ^9^ As a result, polygenic scores for diabetes have been developed to better capture this polygenic risk. ^10^

As for most cancers, persons with a first-degree family history of pancreatic cancer have about twice the risk of pancreatic cancer. ^11^ This increased risk is attributed to inherited genetic factors, lifestyle risk factors, and their interactions. ^5^ There is clear evidence linking familial cases of pancreatic ductal adenocarcinoma to mutations in critical genes, ^12, 13,14^ and mendelian segregation analysis further supports the hereditary aspects of the disease. ^15^ Genome-wide association studies (GWAS) have also identified numerous common single nucleotide polymorphisms (SNPs) that contribute to increased susceptibility. ^16^ Obesity, smoking, and heavy alcohol drinking are risk factors, ^17^ and these may interact with genes to increase pancreatic cancer risk. ^18^

Whether diabetes causes pancreatic cancer was debated for many years, although now consensus for a causal association is being reached. ^15,19^ Several approaches have been used to attempt to distinguish between diabetes as a cause versus the result of pancreatic cancer, including modeling longer versus shorter time between diabetes onset and pancreatic cancer diagnosis, and using mendelian randomization (MR) with genetic instruments for diabetes to evaluate the association with pancreatic cancer. For example, diabetes mellitus is a recognized feature of pancreatic cancer, ^20, 21^ with new-onset diabetes (diagnosed within the past two years) being particularly common among pancreatic cancer patients. ^22, 23^ Previous MR studies, which use genetic data to assess causality, have produced mixed results: some suggest a causal relationship, while others do not. ^24^, ^25^ The inconsistency may be due to limitations in the genetic instruments used, which can weaken the statistical power of these studies. However, more recent MR studies have demonstrated a more consistent positive association, suggesting a strengthening body of evidence in support of causality. ^26^

Distinguishing between new-onset diabetes resulting from yet undetected pancreatic cancer and diabetes as a risk factor for pancreatic cancer is essential for public health action. The former may provide information to improve earlier detection of pancreatic cancer. ^16, 15^ The importance of implementing a screening program for the early detection of pancreatic cancer is critical due to its deadly nature. However, because pancreatic cancer is rare in the general population, a broad screening initiative is not feasible. Instead, it has proposed that focusing on targeted screening for high-risk individuals would be more effective in reducing the incidence of this cancer, ^27–29^ a strategy that is being tested in pilot study in the United Kingdom’s National Health Service. ^30,31^ The latter may provide information to support pancreatic cancer prevention. Preventing disease by intervening on risk factors is a strategy that avoids morbidity and reduces healthcare costs. ^32^ Thus, being able to determine whether diabetes is also a cause of pancreatic cancer is essential evidence for 1) including in diabetes prevention and treatment guidelines the avoidance of pancreatic cancer as benefit, and possibly for 2) testing whether targeting persons with a family history of pancreatic cancer for more extensive screening for diabetes and diabetes management would reduce their subsequent risk of pancreatic cancer.

This study aimed to determine whether genetic risk of diabetes, as measured by a polygenic score (PGS), is associated with a family history of pancreatic cancer in persons without a personal history of pancreatic cancer. If diabetes mellitus causes pancreatic cancer, then we hypothesized that individuals with a first-degree family history of pancreatic cancer, reflecting both inheritance and lifestyle risk factors, are more likely to be enriched with genetic propensity to diabetes than individuals without a first-degree family history of pancreatic cancer. Our hypothesis assumes that the major causal genes for diabetes and pancreatic cancer are not identical.

## Methods

### Study design and population

We conducted a cross-sectional study among 3,911 participants using data from *All of Us*. The *All of Us* Research Program is a National Institutes of Health (NIH) initiative to improve medical precision and population health.^33^ The goal is to enroll about one million participants with the majority coming from historically underrepresented in biomedical research. *All of Us* incorporate electronic health records, surveys, physical measurements, biospecimen (e.g. blood and urine), survey responses, data from wearable devices. The diverse dataset enables researchers to study the interplay between genetics, environment, and lifestyle on health outcomes.^33,34^

*All of Us* is Institutional Review Board (IRB)-approved,^35^ all participants provided informed consent. For this study, we analyzed de-identified data through the secure *All of Us* Researcher Workbench without direct access to raw participant data. This platform provides a controlled environment for data analysis while maintaining participant privacy and security.^35,36^

Figure 1 shows inclusions and exclusions. We included adults aged 30 years or older in the study. The minimum age range helped ensure that we could observe individuals with a family history of pancreatic cancer, as the mean age of pancreatic cancer diagnosis is 70 years. We excluded individuals with a personal history of pancreatic cancer or a known diagnosis of type 1 diabetes or genetic syndromes that predispose to both pancreatic cancer and diabetes (e.g., hereditary pancreatitis, cystic fibrosis).

**Figure 1.**
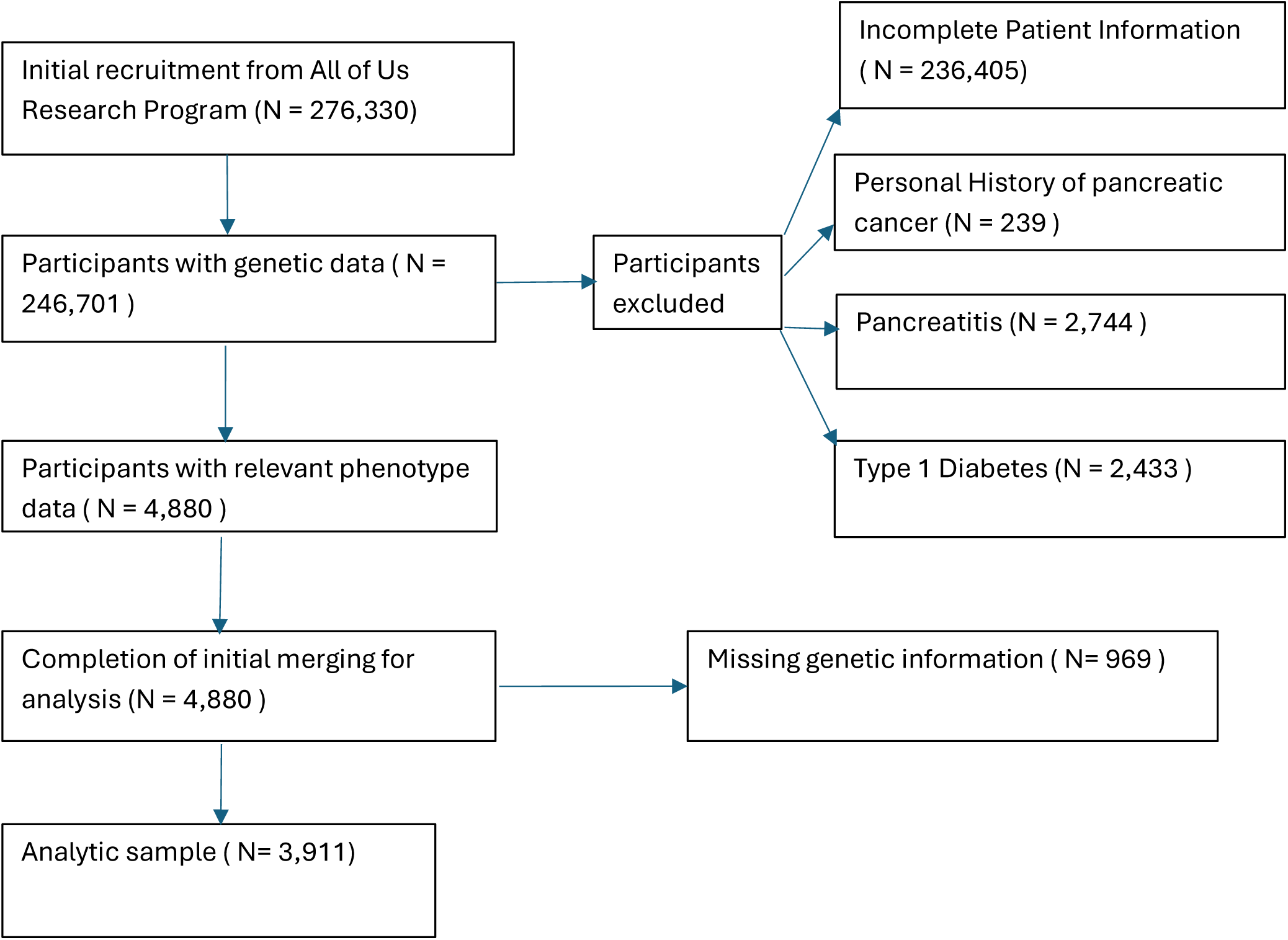
Inclusions and Exclusions, cross-sectional study of 3,911 participants in *All of Us*.

### Assessment of first-degree family history of pancreatic cancer

Family history of pancreatic cancer was assessed in *All of Us* through structured electronic health record (EHR) reviews and self-reported questionnaires capturing first degree relatives’ cancer histories. Participants completed the personal and family health history survey, indicating whether any first-degree biological relatives had been diagnosed with pancreatic cancer. Survey responses were mapped using OMOP PPI vocabulary codes. EHR derived data were assessed using ICD 10 Z80.0 (“family history of malignant neoplasm of digestive organs’’). A positive family history was defined as pancreatic cancer reported in either data source.

### Calculation of a polygenic score (PGS) for type 2 diabetes

The PGS for type II diabetes was calculated using pre-trained weights based on publicly available summary statistics (i.e., discovery GWAS) from the PGS Catalog.^37^ These summary statistics are based on ∼6.9 million SNPs and developed in European-ancestry populations (n = 120,280 for training; n = 159,208 for GWAS discovery). It explains ∼3% of variance in diabetes risk and shows good predictive performance (AUROC ∼0.73) in European samples. Using the short-read whole genome sequencing (srWGS) data in *All of Us*, the data were subset to those SNPs included in the discovery GWAS, and scoring was conducted using Plink 2.0.^38^ The resulting PGS was then regressed on the top 10 genetic ancestry principal components (provided by *All of Us*) and standardized prior to analysis.

### Statistical analysis

Means and prevalences of baseline demographic and clinical characteristics of the study participants, stratified by family history of pancreatic cancer, were calculated. ^33^ Logistic regression models were used to estimate the odds ratio (OR) of family history of pancreatic cancer associated with a high diabetes PGS, before and after adjusting for factors that are associated with diabetes and/or pancreatic cancer.

### Assessing Bias

To explore potential sources of bias, we performed sensitivity analyses and simulations. We examined the effect of excluding participants with a personal history of pancreatic cancer, recognizing that such individuals may have been underrepresented due to illness or mortality. Additionally, we evaluated the potential for survival bias by stratifying the analysis by age.

## Results

High PGS was defined as at or above the 75^th^ percentile. **Table 1** shows the demographic and lifestyle factors of the 3,911 participants included in the analysis. Of them, 173 (4.4%) had a first-degree family history of pancreatic cancer, and 3,738 participants did not. The diabetes PGS is standardized to a mean of 0. Mean PGS was close to 0 in both those with and without a family history of pancreatic cancer. Although the prevalence of daily alcohol drinking was higher in those with family history of pancreatic cancer (**Table 1)**, these individuals had a slightly lower mean diabetes PGS (mean −0.07, standard deviation 0.48, range -1.54 to 1.14) compared to those without a family history (mean 0.003, standard deviation 0.50, range -1.82 to 1.93), and their range was narrower. Of those with a family history of pancreatic cancer, 18.5% (32 of 173) had a high PGS, compared to 25.3% (946 of 3,738) of those without a family history.

**Table 1.** Characteristics of the participants aged ≥30 years old without a personal history of pancreatic cancer, *All of Us*.

| **Covariates** | **Overall (N=3911)** | **With a Family History of pancreatic cancer (N=173)** | **Without a Family History (N=3738)** |
| --- | --- | --- | --- |
| **Age/ years** | Mean 65.7 (30.8-85.8) | 65.58 | 67.40 |
| **Race** |  |  |  |
| White | 83.38 % | 83.41 % | 83.38 % |
| Other (Black, Asian, etc.) | 16.62 % | 16.59 % | 16.62 % |
| **Ethnicity** |  |  |  |
| Hispanic or Latino | 1.33 % | 0.87 % | 1.35 % |
| Non-Hispanic or Non-Latino | 98.67 % | 99.13 % | 98.65 % |
| **Sex at Birth** |  |  |  |
| Male | 35.25 % | 35.37 % | 35.24 % |
| Female | 64.75 % | 64.63 % | 64.76 % |
| **BMI/kg/m<sup>2</sup>** | Mean 34.56 (19-70) |  |  |
| Obese ($\geq 30$ kg/m<sup>2</sup>) | 73.18 % | 71.62 % | 73.25 % |
| Non-Obese ($< 30$ kg/m<sup>2</sup>) | 26.82 % | 28.38 % | 26.75 % |
| **Smoking** |  |  |  |
| Former/ Current smokers | 20.78 % | 20.09 % | 20.81 % |
| Not at all | 79.22 % | 79.91 % | 79.19 % |
| **Alcohol** |  |  |  |
| Never | 74.06 % | 59.39 % | 74.78 % |
| Occasionally | 18.07 % | 12.23 % | 18.36 % |
| Everyday | 7.87 % | 28.38 % | 6.86 % |

In both unadjusted and adjusted models- first controlling for demographic variables, and then additionally for lifestyle risk factors for pancreatic cancer (multivariable-adjusted model), the quartile analysis showed a non-linear association between PGS and family history of pancreatic cancer. Participants in the highest diabetes PGS quartile (Q4) had a lower odd of having a family history of pancreatic cancer compared to those in the lowest quartile (Q1) (multivariable-adjusted OR = 0.66, 95% CI: 0.41–1.04; **Table 2**). In contrast, no significant association was observed for the second (Q2) or third (Q3) quartiles of the diabetes PGS were not associated with family history of pancreatic cancer (**Table 2**). Alcohol drinking did not confound the association between diabetes PGS and family history of pancreatic cancer.

**Table 2.** Association between quartiles of diabetes PGS and family history of pancreatic cancer, *All of Us*.

| Quartile of diabetes PGS | Number of participants with and without a family history of pancreatic cancer | OR | 95% CI | p-value |
| --- | --- | --- | --- | --- |
| ***Unadjusted*** |  |  |  |  |
| 1 | 47/931 | 1.00 | Ref | - |
| 2 | 45/933 | 0.96 | 0.63 - 1.45 | 0.83 |
| 3 | 49/929 | 1.04 | 0.69 - 1.58 | 0.83 |
| 4 | 32/945 | 0.67 | 0.42 - 1.06 | 0.09 |
| ***Adjusted for age, sex, race, and ethnicity*** |  |  |  |  |
| 1 | 47/931 | 1.00 | ref | - |
| 2 | 45/933 | 0.96 | 0.63 - 1.46 | 0.85 |
| 3 | 49/929 | 1.06 | 0.70 - 1.61 | 0.78 |
| 4 | 32/945 | 0.68 | 0.43 - 1.08 | 0.11 |
| ***Adjusted for age, sex, race, and ethnicity, BMI, alcohol, and smoking*** |  |  |  |  |
| 1 | 47/931 | 1.00 | ref | - |
| 2 | 45/933 | 0.96 | 0.62 - 1.46 | 0.83 |
| 3 | 49/929 | 1.01 | 0.67 - 1.54 | 0.94 |
| 4 | 32/945 | 0.66 | 0.41 - 1.04 | 0.08 |

Given this non-linear pattern of association, we next modeled the association of the top quartile (≥75^th^ percentile), which we considered to be higher genetic risk of diabetes, versus the bottom three quartiles (<75^th^ percentile), which we considered to be lower genetic risk of diabetes (**Table 3**). In the unadjusted model, higher diabetes PGS was statistically significantly inversely associated with family history of pancreatic cancer (p<0.04).

**Table 3.** Unadjusted and adjusted odds ratio (OR) and 95% CI of family history of pancreatic cancer for higher (≥75th percentile) versus lower (<75^th^ percentile) diabetes PGS, *All of Us*.

|  | Unadjusted |  |  | Multivariable Adjusted* |  |  |
| --- | --- | --- | --- | --- | --- | --- |
|  | OR | CI | p-value | OR | CI | p-value |
| Diabetes PGS | 0.67 | 0.45-0.98 | 0.04 | 0.67 | 0.44-0.97 | 0.04 |

\*Adjusted for age, sex, race, ethnicity, BMI, alcohol drinking status, smoking status.
\*Adjusted for age, sex, race, ethnicity, BMI, alcohol drinking status, smoking status.

Likewise, the OR was similar in the multivariable adjusted model(p<0.04).

### Assessing Bias

Given that the findings were not consistent with the hypothesis, we performed sensitivity analyses and simulations to assess the possibility of bias.

#### Bias due to a pre-defined exclusion

In the analytic cohort, we excluded participants who had a personal history of pancreatic cancer (Figure 1) because we were concerned that some participants with a personal history of pancreatic cancer would already be deceased or too ill to participate in *All of Us*. We had assumed that this exclusion would be non-differential with respect to genetic propensity to diabetes. However, this assumption could be incorrect.

Persons with a personal history of pancreatic cancer may be more likely to have a family history of pancreatic cancer than persons without a personal history of pancreatic cancer. If our hypothesis is true, then those with both a personal and family history of pancreatic cancer would also be even more enriched with a genetic propensity to diabetes, than participants without a personal history but with a family history. Thus, it is possible that we induced selection bias in making this exclusion. To assess this possibility, we included participants with a personal history of pancreatic cancer and re-calculated the OR for higher PGS and family history of pancreatic cancer (unadjusted given the lack of confounding by covariates). We still observed an inverse association (OR=0.74), albeit attenuated, compared with the main analysis excluding those persons (OR=0.67). We also simulated these ORs for the scenario that diabetes PGS is not associated with family history of pancreatic cancer. When including those with a personal history of pancreatic cancer, OR=1 (by design) and when excluding them, OR=0.90. These assessments suggest a modest (∼10%) selection bias resulting from our exclusion of those with a personal history of pancreatic cancer (assuming no survival bias for entry into *All of Us*).

#### Bias due to differential survival

We also considered whether a higher risk of death or illness among persons with both a higher genetic risk of diabetes and a family history of pancreatic cancer (e.g., because they have both diabetes and a personal history of pancreatic cancer) than only one or neither state may have led to their differential underrepresentation among participants in *All of Us*. First, we stratified by age. In younger participants (≤50 years), among whom we expected a lower likelihood of survival bias (e.g., too young, on average, to have developed both diabetes and pancreatic cancer), the association was less inverse (OR=0.75, 95% CI 0.47-1.18). In contrast, the OR was more inverse in older participants (>50 years, OR=0.60, 95% CI 0.38-0.94).

Second, we performed a simulation in which we sequentially increased the proportion of those in the higher PGS/family history of pancreatic cancer category (i.e., including those who did not survive/were too ill to participate) relative to those in the higher PGS/no family history of pancreatic cancer category. As the percentage increased from the observed 0.94% in that category, the OR became less inverse, null, and then became positive (**Table 4**). For example, when the category proportion increased(doubled) to 1.88%, the odds ratio (OR) was 1.39 when individuals with a personal history of pancreatic cancer were excluded. When those with a personal history of pancreatic cancer were included, the OR increased to 1.54. We next reviewed the probability of being diagnosed with pancreatic cancer or dying of it from birth to various ages prior to entry into *All of Us* using Surveillance, Epidemiology and End Results Program data (**Supplement Table 1**). For birth to age 50, the probability of developing or dying of pancreatic cancer is <0.1% ^39^, suggesting that in the ≤50-year-old stratum, the likelihood of survival bias is very small.

**Table 4.** Simulation of bias correction for differential survival of persons with both a higher diabetes PGS and family history of pancreatic cancer prior to enrollment in *All of Us*.

|  |  |  |
| --- | --- | --- |
| Proportion in the category of higher diabetes PGS and family history of pancreatic cancer | OR when including persons with pancreatic cancer who did not survive or were too ill to participate in *All of Us* |  |
|  | Excluding persons who participated in *All of Us* with a personal history of pancreatic cancer | Additionally including persons who participated in *All of Us* with a personal history of pancreatic cancer |
| 0.94% (observed) | 0.67 | 0.74 |
| 1.18% (25% higher) | 0.84 | 0.93 |
| 1.39% (50% higher) | 1.01 | 1.11 |
| 1.88% (100% higher) | 1.39 | 1.54 |

## Discussion

The relationship between a genetic predisposition to diabetes mellitus and family history of pancreatic cancer likely reflects a complex interplay of shared and distinct risk factors. We hypothesized that if diabetes is a cause of pancreatic cancer, then persons with a family history of pancreatic cancer, reflecting both inheritance and lifestyle risk factors, would be enriched with a genetic propensity for diabetes. Contrary to our hypothesis, persons with a family history of pancreatic cancer were not enriched with a genetic risk of diabetes as defined by a higher diabetes PGS. Through sensitivity analyses and simulations, we conclude that the inverse association between diabetes PGS and family history of pancreatic cancer that we observed may be, in part, due to survival bias. This work points to complexities in the conduct of etiologic research using the cross-sectional design.

Prior studies reported a positive association between a family history of pancreatic cancer and pancreatic cancer risk (OR=2.79 for first-degree relatives), ^40^ while a family history of diabetes is linked to a 30–50% elevated risk of pancreatic cancer. ^41^ However, genetic overlap between diabetes and pancreatic cancer appears limited, as we assumed for our hypothesis. ^38^ Genome-wide association studies have identified distinct susceptibility loci for each condition, with minimal shared genetic architecture. For example, *NR5A2* and *PDX1* are implicated in both diseases, but were not included in standard diabetes PGS. ^42,43^

These data from *All of Us* suggest that a genetic propensity for diabetes, as measured by PGS, is not positively associated with a family history of pancreatic cancer. This contrasts with epidemiological positive links between diabetes and pancreatic cancer but possibly aligns with genetic studies showing limited overlap in susceptibility loci.

We considered the possibility of non-causal explanations for the findings. In the Results, we considered in detail two possible sources of selection bias. With respect to the exclusion of those with a personal history of pancreatic cancer, we estimated that selection bias accounted for about ∼10% of the relative difference from the unbiased OR (assuming no survival bias). With respect to survival bias, we determined that differential survival is very unlikely to account fully for the inverse association in participants ≤50 years old, given the rarity of pancreatic cancer in that age group. Additionally, lifestyle factors (e.g., smoking, alcohol use) and comorbidities like obesity may contribute to familial clustering of pancreatic cancer in addition to genetic predisposition. Indeed, the covariate, alcohol drinking, was positively associated with family history of pancreatic cancer. However, alcohol was not a confounder of the diabetes PGS and family history of pancreatic cancer association, and thus, did not explain the observed inverse association. Furthermore, given the cross-sectional design and study population age distribution, incomplete ascertainment of family history of pancreatic cancer is possible leading to misclassification. We restricted the analysis to a minimum age of 30 to increase the opportunity for the observation of a family history. However, we cannot rule out differences in age of onset of pancreatic cancer in family members based on the genetic propensity to diabetes leading to differential opportunity to ascertain family history of pancreatic at younger ages in the analytic cohort. Pleiotropy could account for the inverse association if the same gene positively influences one condition (e.g., diabetes) and inversely influences the other (e.g., pancreatic cancer). However, the diabetes PGS we used captures common variants (e.g., in *TCF7L2*, *PPARG*) and does not include rare, high-penetrance genes (e.g., *BRCA2*, *PALB2*) linked to pancreatic cancer,^44^ and genes influencing pancreatic development (e.g., *PDX1*) may contribute to both conditions were not included in diabetes PGS.^45,46^ Moreover, it is possible that genetic propensity to diabetes without the development of diabetes (e.g., do not have lifestyle risk factors that may interact with genetic propensity) is insufficient for the development of pancreatic cancer (in relatives). Also, diabetes can develop in persons with a low genetic propensity to diabetes due to the presence of lifestyle risk factors, which could be sufficient to lead to the development of pancreatic cancer (in relatives). The relative balance of these scenarios could lead to inverse, null, or positive associations. Finally, as with any observational study, we cannot rule out chance as an explanation for the inverse association observed.

## Conclusion

Our hypothesis that a genetic propensity to diabetes would be positively associated with a family history of pancreatic cancer, if diabetes is a cause of pancreatic cancer, was not supported in this cross-sectional study in *All of Us*. We explored non-causal explanations for the observed inverse association and determined that selection bias, in part, explanatory. Our work highlights the inferential limitations of cross-sectional studies, even when embedded in state-of-the-art epidemiologic databases. Future research should address these biases through prospective designs or simulation-based approaches.

## Data Availability

Data used in this study are available to registered researchers through the All of Us Researcher Workbench (https://www.researchallofus.org/) following institutional registration and required training.

## Acknowledgements

This research was conducted using data from the All of Us Research Program, a program supported by the National Institutes of Health. The All of Us Research Program is supported by the NIH Office of the Director and other NIH Institutes and Centers under Award Number OT2OD035580. The content is solely the responsibility of the authors and does not necessarily represent the official views of the National Institutes of Health or the All of Us Research Program. All of Us Research Program. All of Us Researcher Workbench. Version [All of Us Controlled Tier Dataset v7]; [access date - 24 January 2025]. https://www.researchallofus.org/

---

**Supplement Table 1.** Cumulative risk of pancreatic cancer diagnosis and death from birth to specified age, US SEER Program*.

|  | Probability of pancreatic cancer event by specified age |  |
| --- | --- | --- |
| Birth to specified age | Diagnosis | Death |
| 30 | <0.1% | <0.1% |
| 40 | <0.1% | <0.1% |
| 50 | 0.1% | <0.1% |
| 60 | 0.2% | 0.1% |
| 70 | 0.6% | 0.4% |
| 75 | 0.8% | 0.6% |
| 80 | 1.1% | 0.8% |
| 85 | 1.3% | 1.0% |
| *Adapted from SEER for 2019, 2021-2022 <sup>39</sup> |  |  |

## Notes

### Competing Interest Statement

The authors have declared no competing interest.

### Funding Statement

This study did not receive any funding.

### Author Declarations

Institutional Review Board of the Johns Hopkins Bloomberg School of Public Health waived ethical approval for this work.

